# Clinical-Grade Validation of an Autofluorescence Virtual Staining System with Human Experts and a Deep Learning System for Prostate Cancer

**DOI:** 10.1101/2024.03.27.24304447

**Authors:** Pok Fai Wong, Carson McNeil, Yang Wang, Jack Paparian, Charles Santori, Michael Gutierrez, Andrew Homyk, Kunal Nagpal, Tiam Jaroensri, Ellery Wulczyn, David F. Steiner, Po-Hsuan Cameron Chen, Luke Restorick, Jonathan Roy, Peter Cimermancic

## Abstract

The tissue diagnosis of adenocarcinoma and intraductal carcinoma of the prostate (IDC-P) includes Gleason grading of tumor morphology on the hematoxylin and eosin (H&E) stain, and immunohistochemistry (IHC) markers on the PIN-4 stain (CK5/6, P63, AMACR). In this work, we create an automated system for producing both virtual H&E and PIN-4 IHC stains from unstained prostate tissue using a high-throughput multispectral fluorescence microscope and artificial intelligence & machine learning. We demonstrate that the virtual stainer models can produce high-quality images suitable for diagnosis by genitourinary pathologists. Specifically, we validate our system through extensive human review and computational analysis, using a previously-validated Gleason scoring model, and an expert panel, on a large dataset of test slides. This study extends our previous work on virtual staining from autofluorescence, demonstrates the clinical utility of this technology for prostate cancer, and exemplifies a rigorous standard of qualitative and quantitative evaluation for digital pathology.

## Introduction

Prostate cancer is the second leading cause of cancer death for men.[44] As for many cancer types, the tissue diagnosis and treatment planning require biopsies, histochemical stains, and pathologic evaluation. In recent years, there has been an enormous interest in the use of Artificial Intelligence, and in particular deep neural networks, in human pathology and in Prostate Cancer specifically. Much work focuses on the use of segmentation and classification models to aid in the diagnosis of histopathology images, and establishes that DNNs can near-match or augment human pathologist evaluation. [13, 16, 17, 18, 19] Another use of AI in the pathology diagnostic pipeline is virtual staining.[1, 2, 3, 4, 6, 7] Virtual staining is the artificial synthesis or prediction of a histochemically stained microscopy image from another image, such as another tissue stain, or from autofluorescence of unstained tissue. Virtual staining is a digital technique that can optimize image data generation and conserve tissue specimens. The combination of virtual staining with deep learning diagnostic models when applied to prostate cancer could improve diagnostic consistency and save time in the patient journey. [7] However, given that virtually-stained images are created by generative AI, the technology requires extensive evaluation to establish clinical validity.

In this work, we use a custom-built high-throughput multispectral fluorescence microscope to scan a dataset of 799 slides (796 unique cases). Serial (or near serial) slides separately underwent H&E and PIN-4 IHC staining, and we trained two deep learning models to predict those stains from the autofluorescence of unstained prostate tissue. We then evaluate the virtual stains using both a reader study with a panel of 12 genitourinary pathologists, and specific computational metrics. See Supplementary Table 1 for dataset split breakdown.

### Prostate Cancer

#### Gleason grading

The focus of many segmentation networks for prostate cancer is Gleason grading. The International Society of Urological Pathology (ISUP) grade group (GG) system[10] is a 5-tier prognostic classification based on the modified Gleason scores (GS) and is used in the 2022 World Health Organization classification of prostate tumors[12]. Briefly, GG1 is GS of 6, GG2 is GS 3+4=7, GG3 is GS 4+3=7, GG4 is GS of 8, and GG5 is GS of 9 and 10. These qualitative grades inform treatment planning and are representative of increasing disease severity.

#### IDC-P

Intraductal carcinoma of the prostate (IDC-P) is a recently defined entity associated with poor prognosis even though it is not a part of Gleason grading.[34] IDC-P is characterized by preservation of basal cells, and malignant epithelial cells in prostatic acinar ducts forming cribriform or micropapillary patterns with nuclear atypia or non-focal comedonecrosis. The reproducibility of IDC-P diagnosis including challenges in distinguishing between IDC-P and high-grade prostatic intraepithelial neoplasia (PIN) are current areas of research.

### PIN-4 IHC

PIN-4 immunohistochemistry (IHC) is a 2-color stain that is widely used in conjunction with H&E in the evaluation of prostate biopsies. In PIN-4 IHC, basal cells are labeled in brown by expression of cytoplasmic CK5/6 and nuclear P63, whereas tumor cells are labeled in red by expression of cytoplasmic AMACR.[35] In prostate cancer diagnosis, PIN-4 IHC in combination with H&E may aid IDC-P detection by pathologists.

## Methods

### Data generation

#### Biospecimens and histopathology

All tissue was purchased from an independent pathology lab, and approved for research use. In this study, the grade group distribution of the prostate biopsies was 19% benign, 25% GG1, 19% GG2, 16% GG3, 9% GG4, 12% GG5; and 20% of all biopsies contained IDC-P. Reports of natural grade group distributions are relatively unbalanced, with high grade groups (GG4, GG5) and IDC-P-positive biopsies each representing <5%. Enriching for less abundant grade groups and IDC-P allows for more comprehensive evaluation across the spectrum of Gleason grades, and sufficient powering of the IDC-P endpoint.

The H&E stain was performed using the Tissue-Tek Prisma Automated Slide Stainer (Sakura Finetek). PIN-4 IHC was performed using the BOND RX autostainer and ChromoPlex 1 Dual Detection reagents (Leica Biosystems) as follows. Tissue sections underwent deparaffinization and rehydration, followed by antigen retrieval in Epitope Retrieval Solution 2 (pH 9). Endogenous peroxidases were neutralized by Peroxide Block. Primary monoclonal antibodies against CK5/6 (1:600; KRT5.6/2438), P63 (1:200; 4A4), and AMACR (1:400; AMACR/2748R) were co-incubated at room temperature for 30 min. Secondary horseradish peroxidase (HRP)-conjugated and alkaline phosphatase (AP)-conjugated antibodies were incubated at room temperature for 8 min and 20 min, respectively. Visualization of targets involved 3,3’-diaminobenzidine (DAB) HRP chromogen for 10 min, DAB Enhancer for 5 min, and Fast Red AP chromogen for 20 min. Tissue sections were counterstained with hematoxylin for 5 min. Finally, stained tissue slides were brightfield imaged using the Aperio GT 450 (Leica Biosystems).

#### Autofluorescence imaging

The imaging system used for obtaining the AF images in this work was a custom-built fluorescence microscope. Slides were loaded using an automated slide loader and scanned using an automated stage. The system uses a low-resolution camera to acquire a widefield image, identify tissue regions, and compute the scanning path. For high-resolution scanning, multi-color excitation LEDs (from ultraviolet to red) were focused through the back of the slide to provide uniform illumination over the microscope field of view. The sample was 2D-scanned to cover the entire specimen. The resulting fluorescence from the sample was collected using a microscope objective (20x, NA=0.8), spectrally split into multiple channels (from violet to near infrared) using dichroic beam splitters, and imaged onto 2D sensors. The raw camera images were combined into a single hyperspectral image after applying corrections including dark-frame subtraction, glass fluorescence background subtraction, ghost image subtraction, flat-field correction, and alignment corrections. The parallelized detection across emission wavelengths combined with the high photon detection efficiency allow for a high signal-noise ratio to be obtained with little photobleaching, and thus the scanning is non-destructive to the tissue.

#### Quality control

We applied multiple stages of quality checks to discard tissues and images to create a clean dataset for training and evaluating the ML models. The first stage excluded slides with insufficient tissue or suboptimal stain quality. The inspection also looked for tissue artifacts such as folds, thick paraffin regions, debris, and contamination with fluorescent dyes. The second stage on autofluorescence scans examined scan characteristics such as focus, intensity uniformity and continuity across traverse boundaries, and tissue clipping. No clinical or identification labels were used in the filtering steps. All QC was based on prior experience in ML for microscopy, and was done before any training of models.

After all the QC stages, we collected a total of 557 pairs of AF-BF images with H&E stains from 556 patients and 602 pairs of AF-BF images with PIN-4 stains from 549 patients.

#### Data splits

Data was split into three sets, in accordance with standard machine learning practice. TRAIN, the training set, was used for the purpose of training the deep neural network. EVAL was used to tune hyperparameters of the network, and for piloting downstream analysis. TEST is the set that was reserved for final validation. All of the results reported in this paper are from TEST, and they are the *only* analysis done on this set as of the writing of this manuscript. More details about the splits can be found in Supplementary Materials.

### Virtual Stainer Construction and Training

The virtual stainer model was a deep learning model inspired by the ‘pix2pix’ paired image-to-image translation approach [7, 15] using a UNet and trained using both conditional and unconditional GAN losses. In the sections that follow, we detail each component of the training architecture, as shown in Figure 4.

#### Co-registration

The physical process of chemical staining and imaging produced changes in the configuration of the tissue sample. Therefore the AF and BF images are not spatially aligned. We implemented a co-registration process to align the AF and BF images, using an affine transform. This process estimated the affine transform in 2 separate stages. 1) First it co-registered both images at low magnification using their tissue masks. 2) It then iteratively refined the initial transform by sampling several points from both images at higher magnifications. Relative shifts were computed for each of the paired sampled points. These are then used to estimate a global affine transform for that magnification. During the process of training, patches with low cross correlation scores are discarded with some probability.

#### Input Normalization

We used all of the 20 channels from our hyperspectral microscope instrument. Next the pair-aligned AF and BF stained gigapixel images were cut up into pairs of patches of size 128 x 128 pixels. Thus the input to the Virtual Stainer model was an AF image of shape 128 x 128 x 20 while the expected output is a BF image of shape 128 x 128 x 3. To account for local errors in the coarse alignment between the AF and BF, we added a 16 pixel padding to each side of the BF images to make them 160 x 160 pixel images. During training, we applied a shift invariance loss (described below) to estimate the precise matching loss between the AF and the padded BF images.

Next, the AF images are normalized to range [0, 1] from range [0, uint16.max] and BF images are normalized to [-1, 1] from range [0, 255].

#### Virtual Stain Generator

The core of the virtual stainer was a UNet neural network which takes a 128 x 128 x 20 patch of AF image and returns the corresponding 128 x 128 x 3 patch of BF stained image (Figure X). The UNet used 5 downsampling and 5 upsampling convolutional blocks. Each convolutional block is composed of a convolution, batch normalization, dropout, followed by a convolution and a batch normalization. In the downsampling blocks, each convolutional block was followed by a downsampling 1 x 1 convolution with stride 2 to reduce the size of the featuremap while doubling the number of channels. The dimension of the embedding created after the encoder was 4 x 4 x 1024. In the upsampling blocks, each convolutional block was preceded by a bilinear upsampling and channel mixing convolutional layer. The final output of the UNet was the predicted virtual stain of dimensions 128 x 128 x 3, where the 3 channels correspond to RGB.

#### Shift-invariant regression loss

The generator network was optimized using a composite loss function that incorporated L1 and L2 (MSE) losses on the pixel-wise difference between the generator output and the corresponding ground truth BF images. To account for the inexact co-registration between the AF and BF images, we implement an on-the-fly shift optimization to identify the best fit between 128 x 128 x 3 predicted images and 160 x 160 x 3 label images. Before calculating the regression loss, we first calculated a moving cross-correlation between the predicted image and label image and identify the location of highest correlation. Then the L1 and L2 loss were calculated between the predicted image and 128 x 128 patch of the label image centered at the point of highest correlation, and loss was back-propagated using this shift. This compensated for small (<32 pixel) local deviations in the alignment between AF and BF images.

#### Rotational consistency loss

To make the output rotation invariant and prevent the model from learning an orientation bias, we added a rotational consistency loss. This was implemented as the MSE loss between a rotated virtual stain and the virtual stain produced from a rotated AF image. At each training step, a random rotation, of either 90, 180 or 270 degrees, was applied to the pairs of virtual stains and AF images and the resultant MSE loss was added to the total loss.

#### Unconditional adversarial loss

We trained an unconditional discriminator network that took the real chemically stained BF label images (labeled 0) and virtually stained BF images predicted by the generator (labeled 1) as input. The network is then trained to predict the label i.e. to differentiate between real and virtual stains. The discriminator was trained using MSE between the discriminator’s predictions and the label. This is the classic adversarial loss which increases as the generator gets better at making realistic virtual stains and decreases as the discriminator gets better at differentiating between real and virtual images. Therefore the minmax game between the generator and discriminator forces the generator to produce highly realistic images in order to fool the discriminator. The discriminator neural network was made up of 4 convolutional blocks each consisting of 1 2D convolutional layer, 1 ReLU layer, 1 batch normalization and 1 dropout layer.

#### Conditional adversarial loss

In addition to the unconditional discriminator, we also trained a conditional discriminator which took the concatenated AF and BF image pair as input. During training, the discriminator got two sets of inputs - the first set of inputs used the AF images paired with the corresponding real chemically stained BF images (labeled 0) while the second set used AF images with the corresponding virtually stained BF images predicted by the generator (labeled 1). Again, the discriminator was trained using least squares loss to predict this label (i.e. discriminate between real and virtual BF images), this time, conditioned on the AF image. The discriminator neural network was made up of 4 convolutional blocks each consisting of 1 2D convolutional layer, 1 ReLU layer, 1 batch normalization and 1 dropout layer.

#### Training

We conducted hyperparameter tuning on the evaluation dataset to identify the best hyperparameters. The final set of hyperparameters had dropout at 0.1, batch normalization momentum at 0.8. The convolutional kernel sizes were fixed at 3 x 3 for the generator downsampling blocks, 2 x 2 for generator upsampling blocks, and 4 x 4 for the discriminator blocks. The end-to-end model was trained for 150,000 steps.

### Virtual Stainer Evaluation

The problem of quantitatively evaluating the quality of images produced by generative AI is neither new nor solved [CITE]. However, over time, a variety of techniques and metrics have developed, and it is clear that there is no one-case-fits-all solution. Evaluation metrics can be quantitative or semi-quantitative, general or purpose-fit. We compare the images produced by our Virtual Stainer to ground-truth real images using a variety of different metrics, chosen specifically with the clinic in mind. For both stain types, there is both a human-evaluation and a computational evaluation. See Discussion for further expounding on the virtues of these metrics.

- We compare the automated Gleason Grade given to the real and virtual H&E images, by a purpose-built artificial neural net. The neural net is only trained on real images from a different dataset. Therefore, if it gives the same grades to these virtual and real images, they likely contain similar biological information.
- While a purpose-built artificial grading system like the above is the gold standard of computational analysis for generative images, no such network exists for PIN-4 images. Therefore, for PIN-4, we use a battery of interpretable custom-built computational metrics.
- We compare human Gleason grading of real and virtual H&E images, using standard ORH. (See MRMC)
- We compare human IDC-P diagnosis of real and virtual PIN-4 images, using standard ORH. IDC-P is the most widely accepted semiquantitative clinical measure of PIN-4 stains.

By treating real and virtual images as two different conditions, and demonstrating that diagnostic outcomes from clinicians are non-inferior, we provide a strong validation for the clinical use of these virtual images. To further understand and characterize the degree to which the images contain the same information, we further validate using our battery of computational metrics.

#### Gleason Grading Model for H&E Evaluation

We utilize a machine learning model previously described in [19] to automatically perform Gleason grading. The model was developed on real H&E images that were not included in the datasets used in this study. Automated Gleason grading is performed in two stages: Gleason pattern segmentation, and summarization into core-level grade. Because the training data for the model did not involve the BxChip format, manual annotation was included to segment the biopsy tissue that is used by the summarization algorithm and exclude the non-tissue BxChip region. The output of the model includes benign, GG1, GG2, GG3, GG4-5. The model does not distinguish between GG4 and GG5 due to the original training data groupings, and this grouping is preserved so that the results are comparable to the original publication.

#### Computational Metrics for PIN-4 Evaluation

To evaluate the virtual PIN-4 model, we lack a clinically-relevant AI algorithm. So instead, we used a series of computational metrics. In this case, and in the case of generative image AI in general, it is a non-trivial problem to determine an appropriate metric for image comparison. [5, 36] No metric is perfect, and any metric which has been an optimization target, is suspect of being overfit. Furthermore, if two images are not perfectly identical, how do we determine if the difference is a problem with the image, or a problem with the metric? Finally, there is a degree of natural stain variation in valid staining protocols, as well as potential tissue distortion between staining processes. In [5], the authors discuss many of these issues as they compare virtual staining techniques, and they evaluate how different generic-image comparison metrics stack up against human raters. We include many of these metrics, but also posit that if Virtual Stainer is to be a circumstantial substitute to classic chemical staining protocols, we need to have a sense of how much it differs from real stains, *with respect to relevant biological information*, and whether this amount of variance is expected *within* real stains.

We address this challenge in two ways: First, we use metrics that have some degree of human interpretability, most importantly, Jaccard Distance [33] evaluated on biologically-meaningful segmentations derived using classic color deconvolution, and precision/recall of segmented targets. (See Supplementary Methods) Second, we compare our metrics on a control: PIN-4 staining using serial sections of a prostate tissue microarray (TMAs). In pathological clinical practice, judgements about a case are considered valid on serial sections.[11] We posit that for the practical clinical setting, metrics of differences between serial sections are within what could be considered normal variation. As such, we present metrics for differences between real and virtual prostate patches, and compare these to the same metrics evaluated on serial PIN-4 TMA images. TMAs are good candidates for this use case, as they are constructed to represent a fixed distribution of reference tissues and patients while minimizing wasted tissue.

#### MRMC H&E and PIN-4 Evaluation

Gleason grading is a subjective interpretation with 30-50% interpathologist discordances. However, grading by genitourinary pathologists is more reproducible than general pathologists without subspecialist training. Grading by the majority-vote of clinicians has also been shown to improve reproducibility. As such, to obtain rigorous reference diagnoses for validation, each biopsy was read independently by 2 genitourinary pathologists, and a third to break the tie if the first two disagree. This creates our ‘reference standard’.

The pathologists reviewed a total of 421 TEST cases, in random order with a washout period between real and virtual staining modalities of at least 4 weeks. Each pair is made up of a real, chemically stained BF image and virtually stained image predicted from the AF image of the same tissue sample. Thus for each tissue sample, we obtained 2 sets of grades - one graded on real stains and one scored on virtual stains. In addition to the grades, pathologists have also provided feedback on whether the stain quality met their clinical review standards, almost all of which did, for either condition. (See Supplementary Figures 3 and 4)

For Gleason grading, the subspecialists classified the biopsy as: non-tumor (Benign, ASAP, HGPIN), non-Gleason gradable variant, or Grade Group (GG) 1, GG2, GG3, GG4, or GG5. For intraductal carcinoma of the prostate (IDC-P), the subspecialists indicated whether IDC-P is present on the slide or not, as evaluated from the PIN-4 stained image. As such, the AI and pathologist Gleason Grading proficiency were evaluated against the reference diagnoses as a 6-class classification, while pathologist IDC-P detection was evaluated as a binary (2-class) classification.

Diagnostic data on both real and virtual stains were collected via a multi-reader, multi-case (MRMC) study design (Figure 2). A total of 12 general pathologist readers provided GG and IDC-P classifications as described in the ‘reference standard’.

**Fig 1.**
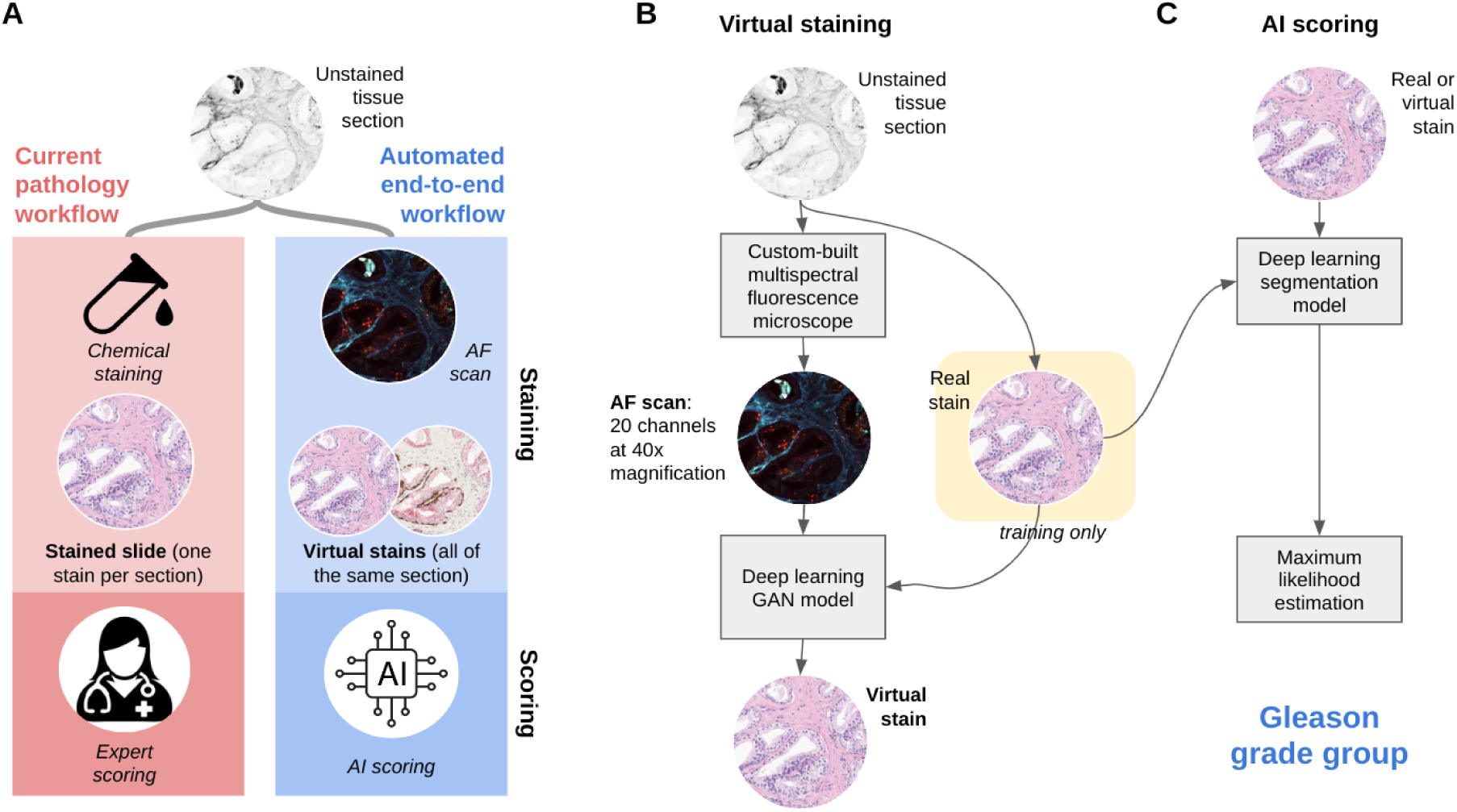
a) We present an automated workflow with virtual staining and AI scoring that mimics the steps of the current pathology workflow. b) The Virtual Staining pipeline uses a custom multispectral microscope to image unstained tissue samples. Our deep learning virtual stainer model takes AF images as input and returns stains images that are trained to look like real chemically stained tissue samples. c) Our AI scoring models use BF images of chemically or virtually stained tissues to generate the Gleason grade group.

**Figure 2.**
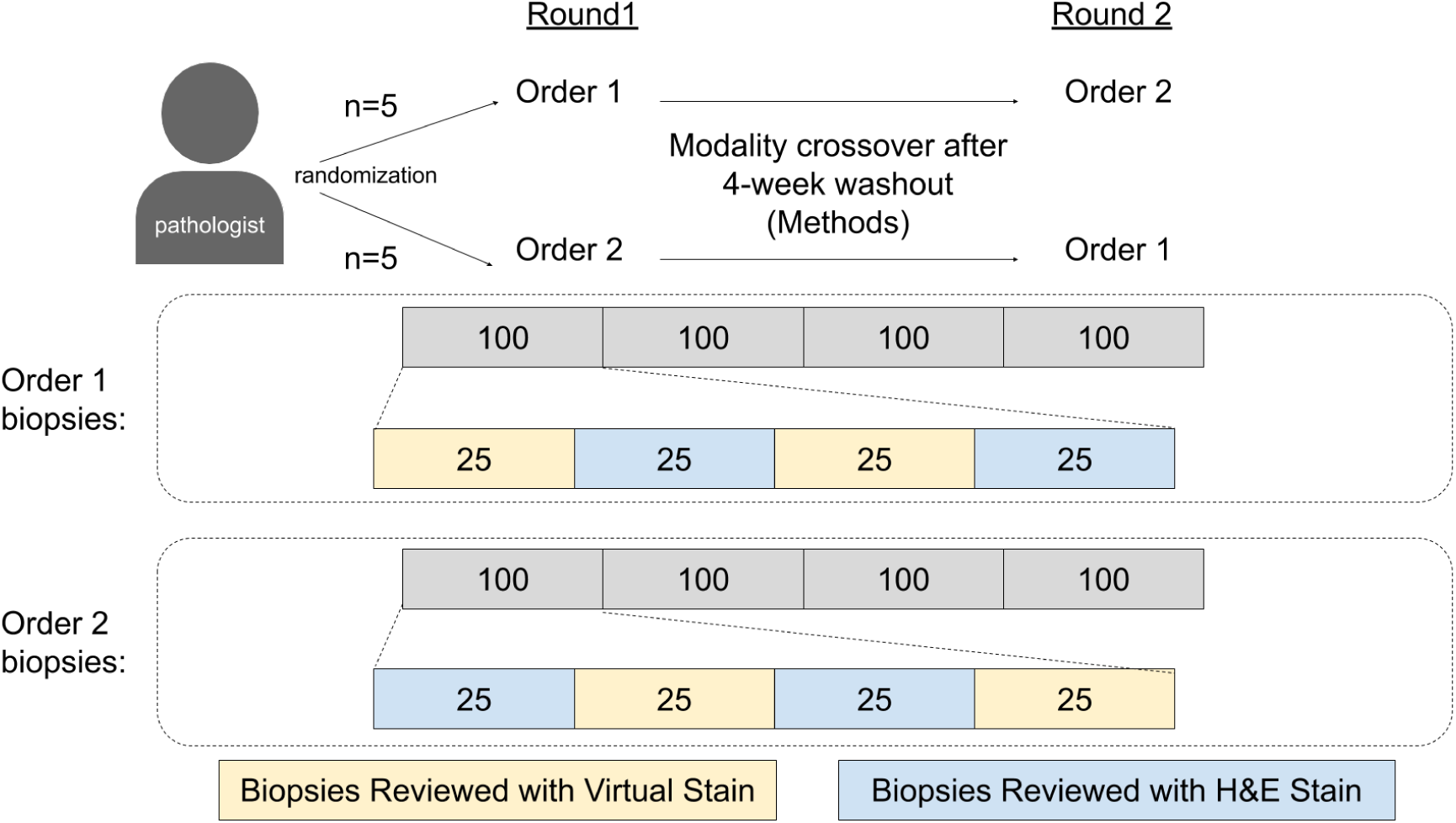
Schematic of proposed multi-reader multi-case study with washout. Proposed methodology for evaluation of both H&E Gleason Grade and PIN-4 IDC reviews by expert pathologists.

The Obuchowski-Rockette-Hillis procedure, which is a standard approach for MRMC studies[32] and accounts for the variance across both readers and cases, was used for non-inferiority (and superiority) testing [30, 31, 32]. We used a non-inferiority margin of 0.1, which is strict for this field [31]. Non-inferiority testing is the accepted standard in pathology for determining if a diagnostic manipulation is clinically valid.

## Results

### Multispectral Microscopy

We built a multispectral fluorescence microscope to probe the near-ultraviolet through the near-infrared portion of the excitation-emission space of the unstained tissue (see Methods section for technical details). To reduce photobleaching and minimize scan time, the microscope uses a parallelized detection scheme to collect 8 wavelength bands simultaneously. The 20 collected excitation-emission combinations provide sufficient spectral resolution to distinguish known autofluorescence features such as collagen, elastin, NADH, flavins, lipopigments, and porphyrins [27]. To illustrate the typical level of spectral variation observed between different types of features, Fig. 3a shows fluorescence spectra, averaged across a single tissue section, computed using the four H&E and PIN-4 stain colors as masks (using real-stained, adjacent sections). Another way to illustrate the spectral information content is to perform linear projections across the spectral axis of the multispectral image. Fig. 3b shows a set of projection vectors used for this purpose, derived from a combination of Canonical Correlation Analysis and Non-Negative Matrix Factorization. Fig. 3e shows a uniform projection (a simple average across all spectral channels) while Figs. 3f-h show spectral projections that enhance stroma and secretory material; more localized extracellular matrix features; and punctate, longer-wavelength fluorescence features often associated with prostatic epithelium. The corresponding H&E and PIN-4 stained regions are shown in Figs. 3c-d. While this simple projection procedure demonstrates the richness of autofluorescence data, it also suggests that a spectral weighting alone cannot recover a sufficiently clean signal to replace staining techniques. A model that can reproduce modern stain methods must incorporate more complex, nonlinear spectral weighting schemes and/or morphological features of the tissue as well.

**Fig. 3.**
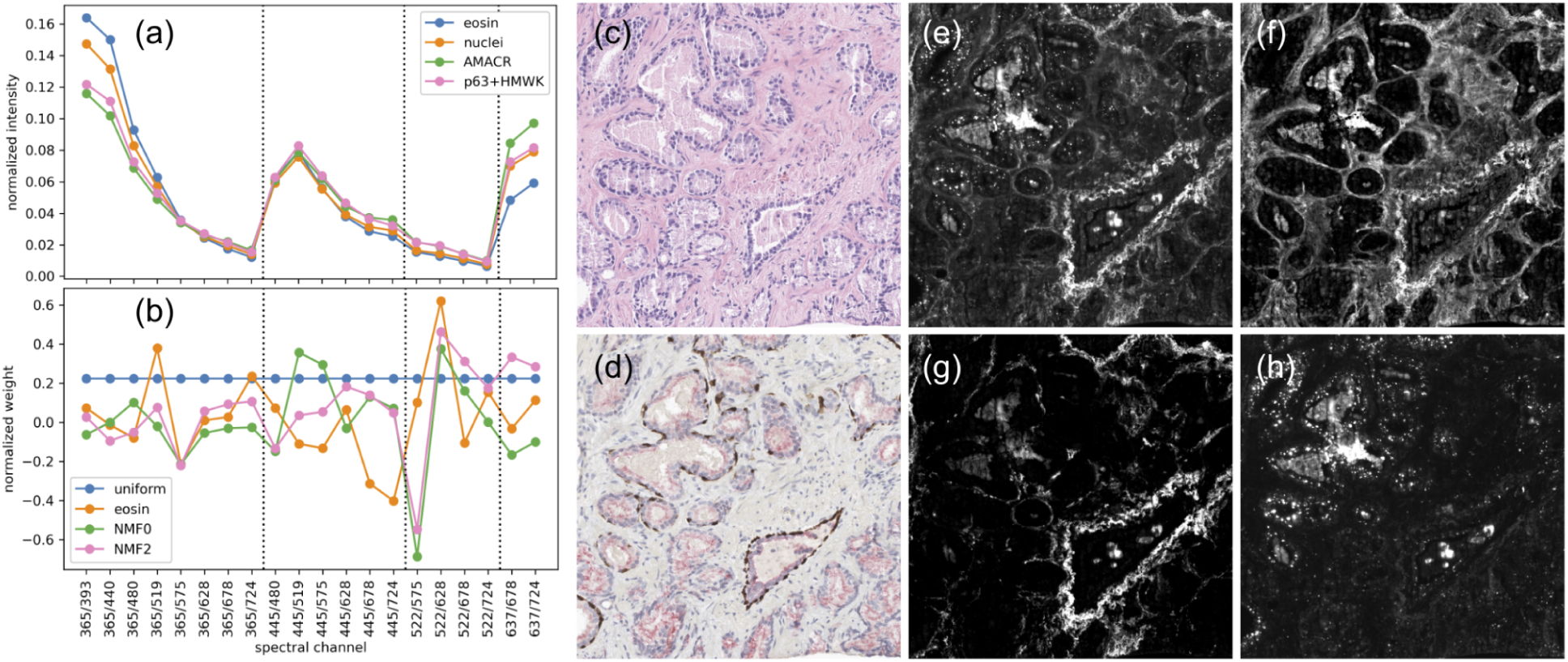
(a) Normalized fluorescence spectra averaged across a whole slide using unmixed, co-aligned and thresholded H&E and PIN-4 images as weights. (b) Weights used to generate the linear spectral projections in e-h. (c) Co-aligned H&E-stained image from the same slide as shown in e-h. (d) Co-aligned PIN-4 from an adjacent section. (e-h) Linear spectral projections of the multispectral fluorescence image using (e) a uniform projection, and projections highlighting (f) eosin features, (g) a subset of extracellular matrix features, and (h) punctate features associated with epithelial regions. [Screenshot taken from [slide](https://docs.google.com/presentation/d/1evbGMqThcmcPqAXhD3bLr-qlxX5_aKp4P4ISrd0dfdg/edit?usp=sharing&resourcekey=0-nmNlry5JAW59ofMfOsTcWQ). Unnormalized spectra could be shown as an alternative. Plots and images generated from [Colab notebook](https://colab.corp.google.com/drive/1zrySbpmg5UvJLvfRxeUoAknvl5H4PewT?resourcekey=0-7iitttqIOSq0Z-t51AyLig&usp=sharing)]

### Virtual Stainer

The virtual stainer model is an image-to-image translation model that takes an autofluorescence image as input and predicts its corresponding virtually stained image. This model is trained using the real chemically stained image as the target output, with a combination of regression and adversarial training objectives (Fig 3, see Methods: Virtual Staining for more details).

The virtual stain models were evaluated using three independent approaches: (1) automated Gleason grading of H&E images, (2) computational comparison of PIN-4 images, and (3) human expert reader studies using Gleason grading and IDC-P assessment.

#### Automated Gleason grading on H&E images

To further demonstrate the VS model’s utility, we evaluate its compatibility with an automatic Gleason grading algorithm [19]. The virtual H&E images were given as an input to the unmodified automatic Gleason grading algorithm that was developed using real H&E images. The algorithm is also applied to the real H&E counterparts of the virtual images. The concordance of the algorithm’s output on real and virtual images is very good, where the quadratic-weighted kappa is 0.902 (95% CI: 0.880, 0.922), and the accuracy is 0.862 (95% CI: 0.847, 0.877). Figure 5a(left) shows the confusion matrix between the output of the algorithm applied to virtual and real H&E images. These numbers are exceedingly high, even for agreement between human raters. Compare to [43]’s human quadratic-kappa of 0.7, or 0.8 with AI-assistance. The high degree of agreement between the automated Gleason grading algorithm output on virtual and real images demonstrates the similarity of the real and virtual images with respect to grading-relevant signals. These results are displayed in Table 1.

**Fig 4.**
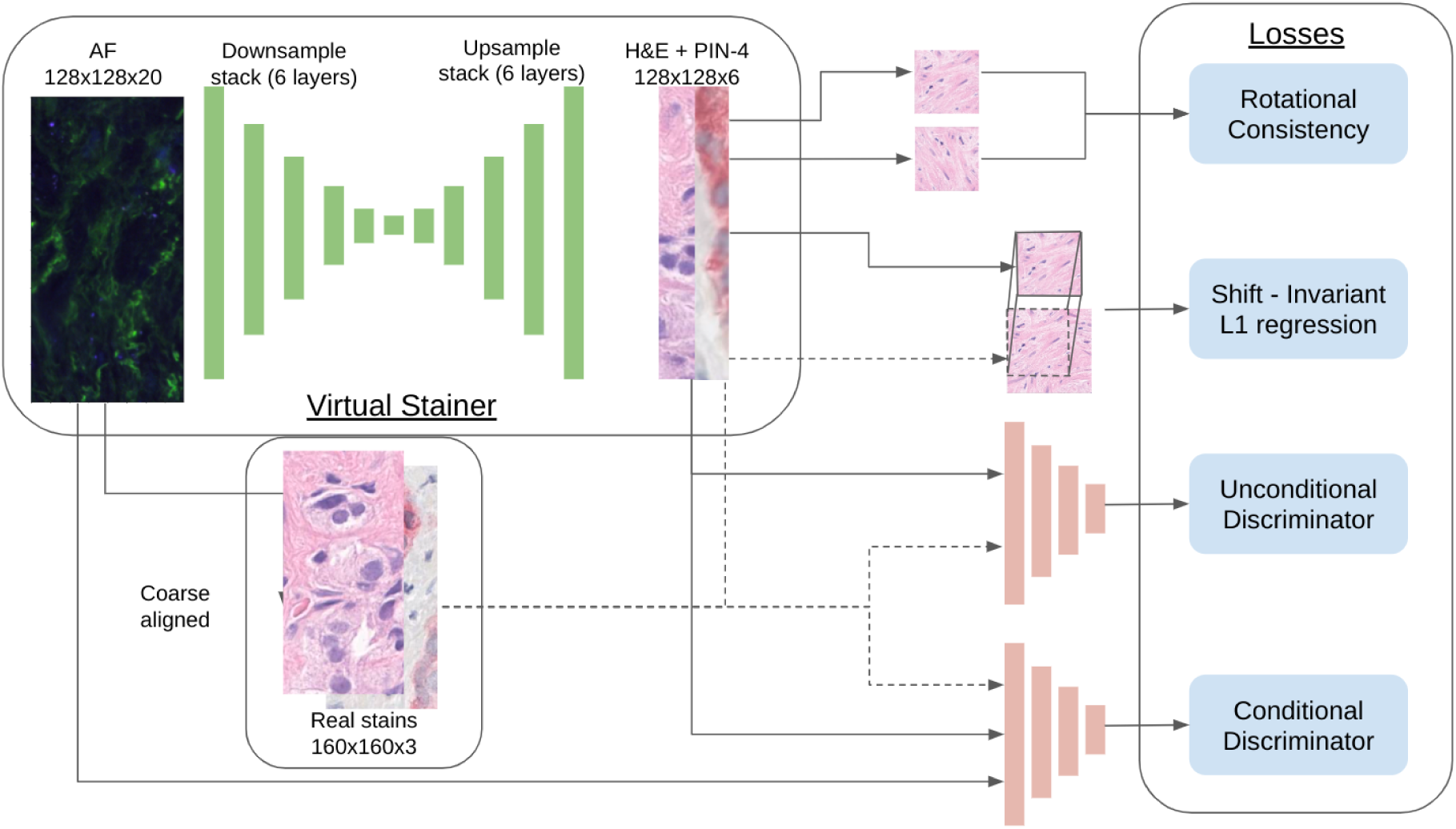
Virtual stainer training pipeline. The virtual stainer is a neural network with a Unet architecture. It takes patches of AF images of size 128×128 and returns 2 images - 1 H&E and 1 PIN-4 BF image. We use chemically stained BF images aligned to the AF images to train the virtual stainer using four different training losses. More details on the architecture and training objectives can be found in the Methods section.

**Fig. 5.**
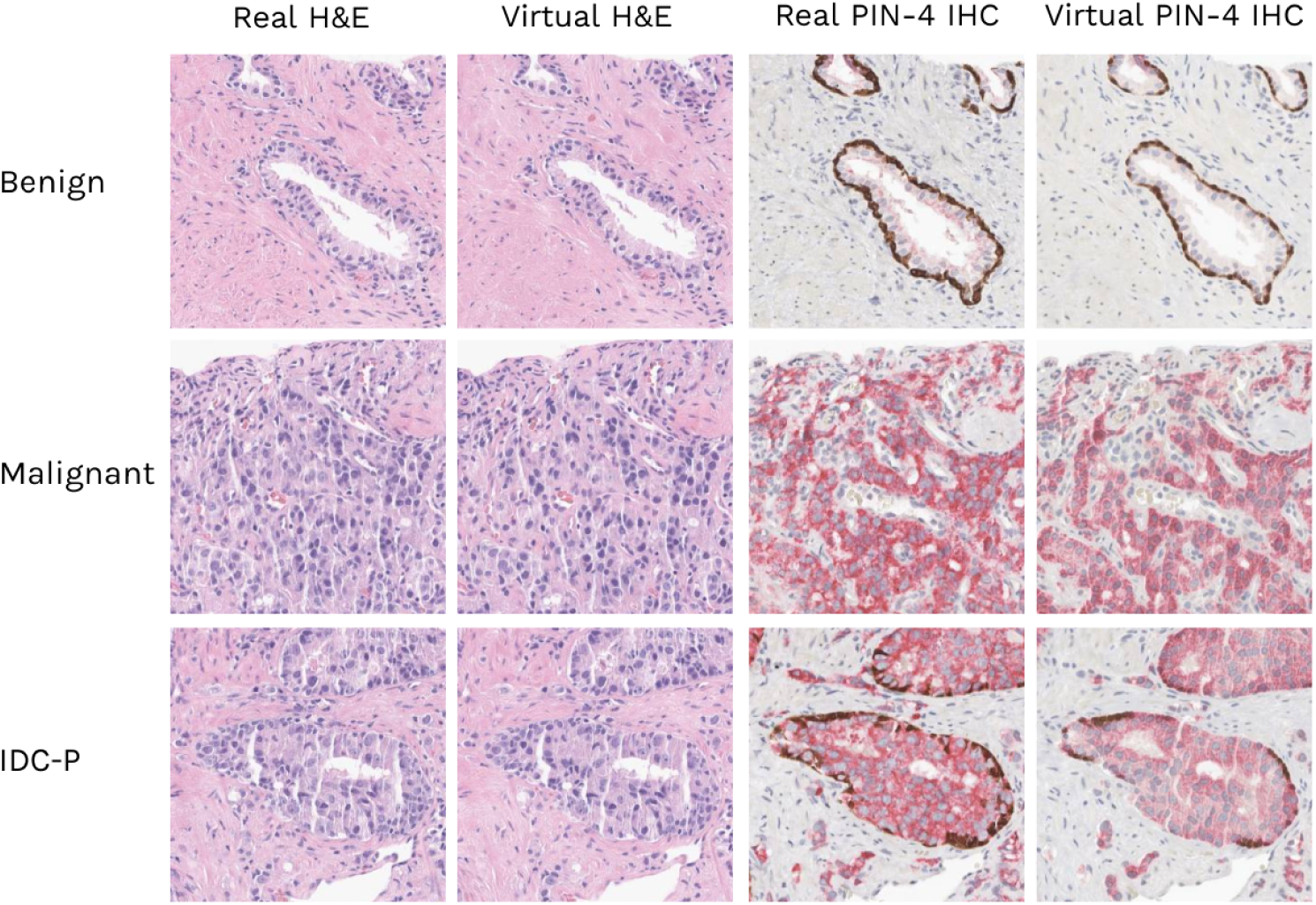
Comparing real and virtual stains at 20X magnification shows that virtual stains accurately capture the morphological attributes of real stains. Here we show examples of chemically stained patches which show benign tissue, malignant tissue, or IDC-P and their corresponding virtual stain.

**Table 1:** Concordance of automatic grading by [19] between Virtual and Real PIN-4 images. The 95% confidence interval for this concordance is given in parentheses, and was obtained by bootstrapping over slides. These values exceed expected human performance substantially [43]; a quadratic-kappa of 0.8 is considered very good.

| Metric | Virtual vs. Real PIN-4 Slides |
| --- | --- |
| Quadratic-Weighted Kappa | 0.902 (0.880, 0.922) |
| Accuracy | 0.862 (0.847, 0.877) |

While the high agreement is encouraging, the algorithm could be suffering from the same error mode both on real and virtual H&E slides, leading to a high concordance. Our trust in the result in Table 1 is therefore bounded by our trust in the algorithm on these slides. While the algorithm is validated in [19], we further demonstrate the performance of the virtually stained H&E by studying the automatic gleason grading model in comparison to human pathologists on this particular set of slides. Ten general pathologists and three genitourinary (GU) specialists were asked to grade one core per slide. The consensus between the GU specialists was taken as the gold standard. We performed Obuchowski-Rockette-Hillis analysis [32] to evaluate the quality of the virtual H&E automatic gleason grading output with a non-inferiority margin of 0.1, which is strict for this field [31]. We found that the output of automatic algorithm is non-inferior to the pathologists (p <0.001). Figure 5b(right) compares the concordance of the model and pathologists with the GU specialists. Our model’s concordance is comparable to the pathologists.

#### Computational analysis of PIN-4 images

We evaluate the Virtual PIN-4 model using a series of computational metrics described in the methods section of this paper. For every one of the metrics, we divided the TEST dataset into patches of uniform size, and compared the real PIN-4 stained patches with the exact Virtually-stained patches to which they correspond. We can see the results of these comparisons in the first column of Table 2. As a basis for comparison, in the second column, we also show metrics for a dataset consisting of serially sectioned TMA slides representing a fixed distribution of PIN-4 stained samples, and where serial sections are considered the smallest unit of slide variation for practical purposes. An example image of these serial TMAs is shown in Supplementary Figure 2. Metrics for TMAs are bootstrapped over TMA sections to provide 95% confidence intervals. Therefore, we consider it useful to see how the difference between real and virtual patches compares to differences between aligned serial TMAs. Patch sizes and magnifications were chosen to match between the two cases. Metrics which are significant are marked with a *.

**Table 2:** Computational Metrics comparing Virtual and Real PIN-4 images. In the first column we can see our battery of comparison metrics applied between real and virtual PIN-4 patches. This is compared to the second column, where we can see the metric between serial PIN-4 TMA sections.

| Metric | Virtual vs. Real PIN-4 Patches | Real PIN-4 TMA, Serial Sections |
| --- | --- | --- |
| AMACR Jaccard Distance* | 0.27 | 0.18 (0.15, 0.19) |
| Nuclear Precision* | 0.93 | 0.87 (0.82, 0.88) |
| Nuclear Recall* | 0.76 | 0.87 (0.84, 0.88) |
| Basal Cell Precision* | 0.39 | 0.17 (0.10, 0.23) |
| Basal Cell Recall* | 0.4 | 0.16 (0.08, 0.17) |
| Average L1 Distance (RGB values) | 13.7 | 11.3 |

As can be seen in the table, every metric aside from Nuclear Recall is significantly better for the Virtual vs. Real patches. This is discussed in the discussion section.

#### Expert reader studies using Gleason grading and IDC-P assessment

We next performed a comparison study to assess the quality of virtual H&E and PIN-4 stains by measuring the difference in Gleason grading and IDC-P assessment, done by pathologists blindly reviewing either chemically stained or virtually stained images.

First, we have defined the ‘reference standard’ Gleason grades and IDC-P statuses, by having 6 senior genitourinary pathologists with 15-30 years of experience reviewing both real and virtual H&E and PIN-4 images. A Gleason grade and IDC-P status for each image was provided by 2 genitourinary pathologists asynchronously, and a third to break the tie if the first two disagreed.

We then performed a multi-reader, multi-case (MRMC) comparison study with 12 general pathologists, as described in the Methods section.

We observed a high agreement between the Gleason grades on real and virtual stains, with the difference in quadratically-weighted kappa values as measured against the ground truth of −0.0016 (95% CI of −0.0193, 0.0161; Table 3, Figure 6). The difference in the kappa value is well above than the study margin of −0.1, suggesting a strong non-inferiority of real versus virtual H&E stains (P-value of 8.4 x 10^-9).

**Figure 6.**
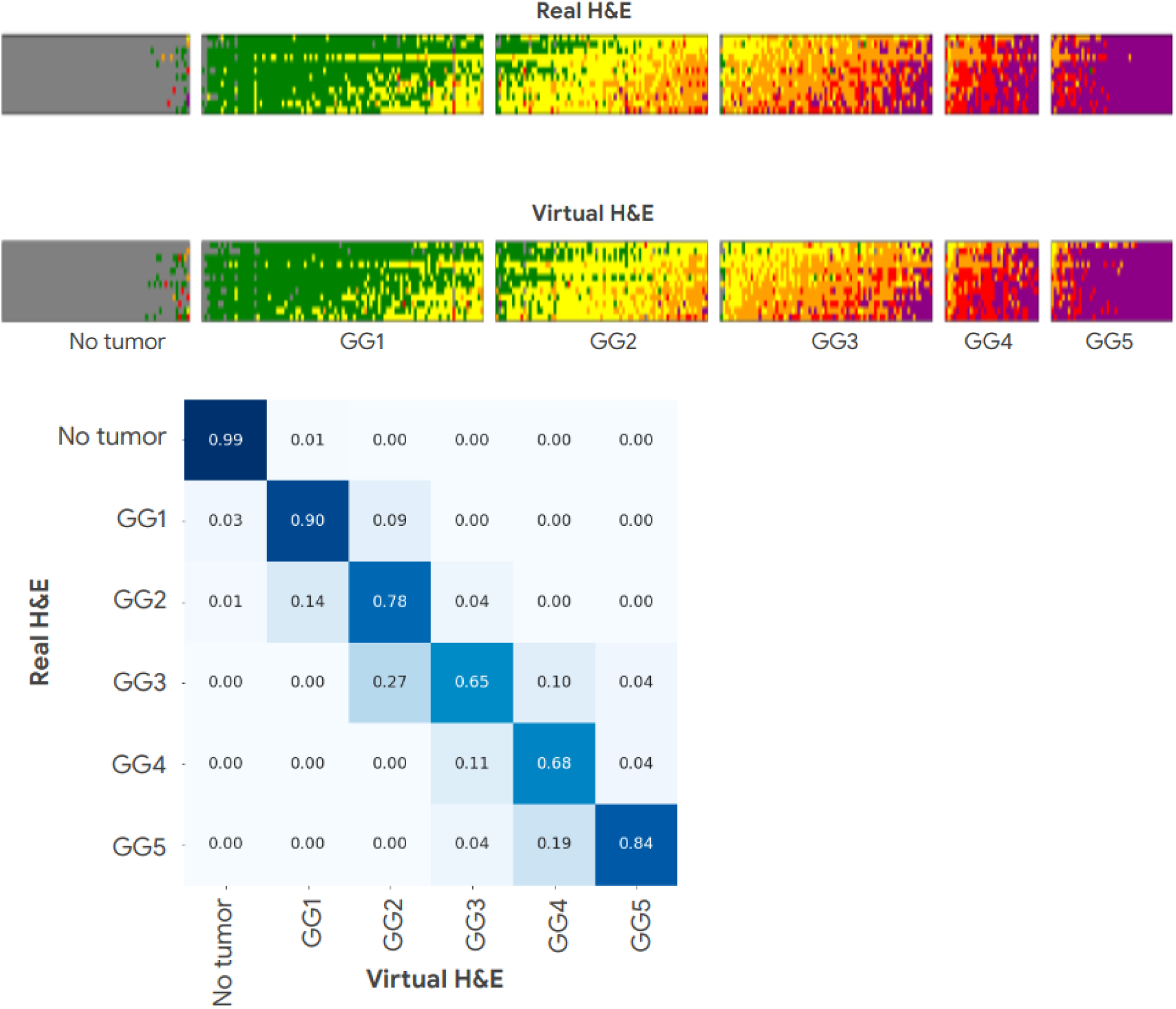
We can visualize human pathologist ratings for both the real and virtually-stained cases. In **A**, we see the full set of 12 raters’ judgements about each case. Each row represents a rater, and each column represents a case, in the same order for both real and virtual conditions. Cases (columns) are divided into each of the 6 subfigures on the basis of their ground truth diagnosis by senior raters. Each case rating is color coded. Grey is No Tumor, Green is Gleason Grade 1, Yellow is GG2, Orange is GG3, Red is GG4, and Purple is GG5. This allows us to observe that raters have consistent patterns of diagnosis between the two conditions even when they differ in opinion from each other. In **B**, we summarize the same data we see in **A** with a confusion matrix. Each cell represents the fraction of cases given a grade in the Real condition that have been graded as each different grade in the Virtual condition. Grades are the median of all 12 raters. Values on the diagonal have been given the same grade in each condition. We can see that this is the case for the vast majority of cases, and not a single case has a disagreement of more than two severity levels. This demonstrates an overall very high level of rater agreement, whether a rater is viewing a case as a real or virtual stain.

Similarly, we observed a high agreement between real and virtual IDC-P status assignments, with the difference in quadratically-weighted kappa values as measured against the ground truth of −0.0925 (95% CI of −0.1382, −0.0469;Table 3, Figure 7).The estimated kappa value between real and virtual is high,, suggesting clinical utility of our virtual PIN-4 stain; however, the non-inferiority result is not statistically significant (p=0.37), and either bigger test set size or further improving the virtual PIN-4 stain will be needed for a more confident claim. That statistical significance could not be achieved with such an exhaustive test set size is reflective of the noisiness of pathologist IDC judgements. The overall performance of pathologists was fairly high (kappa values of 0.69 and 0.60 for real and virtual reads, respectively), but with 3 notable outliers with differences in real kappa values below 0.65 (Figure 7). While the results measure real-world performance of pathologists, this observation suggests that further calibration and training of pathologists for the IDC-P status assignment task could further improve the real vs virtual stain assessment performance.

| | $\Delta\kappa$ | 95% CI | Non-inferiority<br>p-value |
| --- | --- | --- | --- |
| **H&E Real vs. Virtual**<br>**- Gleason Grade** | -0.0016 | ( -0.0193, 0.0161) | $8.4 \times 10^{-9}$ |
| **PIN-4 Real vs.**<br>**Virtual - IDC-P**<br>**Status** | -0.0925 | (-0.1382, -0.0469) | 0.37 |

**Figure 7.**
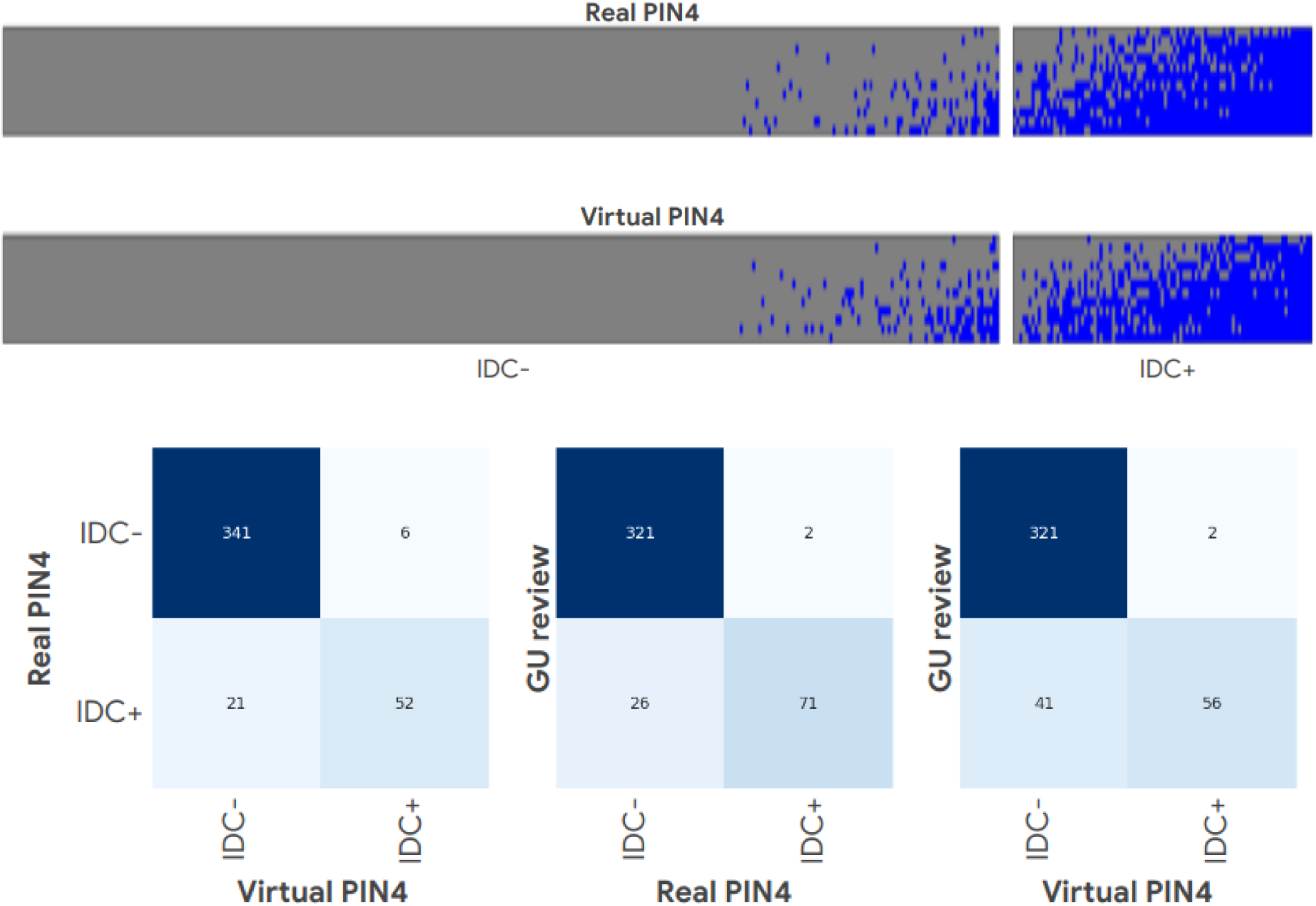
As in Figure 6, in Figure 7 we can see the results of human ratings on real and virtual images. **A** displays the entire dataset of reviews. Each row represents a rater, and each column represents a case, in the same order for both real and virtual conditions. Cases (columns) are divided into each of the 6 subfigures on the basis of their ground truth diagnosis by senior raters. IDC+ ratings are blue, whereas IDC-ratings are grey. **B** shows a summary of **A** as confusion matrices. We can see that median rater agreement for the virtual vs real conditions is similar to rater agreement between median rater and GU expert evaluation. Furthermore, rater agreement on virtual stains is similar to GU expert evaluation on real stains. In all cases, false negatives are substantially more likely than false positives.

We also visualize human rater differences for the real and virtual cases for the PIN-4 stain IDC judgment. In this case, rather than a scale, like Gleason Grade, Intraductal Carcinoma is a simple +/- binary judgment.

## Discussion

In this study, we introduce a Virtual Stainer model that can produce both H&E and PIN-4 IHC stains from autofluorescence of unstained prostate tissue, following our work in [7]. This model is based on a custom autofluorescence imaging platform designed for very high slide throughput and photon efficiency, which enabled us to build the largest-ever (as known to the authors at time of publication) training set for virtual staining based on multispectral autofluorescence.

As new virtual staining results enter the literature[1–6], it becomes increasingly important to establish standards of evaluation. Generative image networks are well known to “hallucinate” outputs [42], and in a clinical use setting, models must be verified to produce diagnostic variance comparable to typical histochemical stains. In addition, we evaluate the model using a series of computational approaches, as well as a human pathologist reader study. To our knowledge, this is the most extensive evaluation using subspecialty pathologists of any virtual stainer model to date, and we hope that it will provide a blueprint for rigorous validation of future virtual stain technologies. We demonstrate that our H&E virtual stainer model is non-inferior to real stains as an input to Gleason grading. This suggests that Virtual Staining could substitute for real stains in that use case.

We further demonstrate the clinical utility of our virtual stainer by creating and evaluating a model that predicts PIN-4 IHC. In a deployed setting, both H&E and PIN-4 stains could be generated from the same AF image of a single tissue section. Because the AF scan is non-destructive, the tissue could then be used for other purposes. We evaluate our virtual stain according to IDC-P status. While the results are clearly comparable, we do not prove non-inferiority (in contrast to Gleason grading), in part due to the known lower reproducibility of pathologist judgment for IDC-P, which is a more clinically challenging entity.

To quantitatively evaluate how our virtual stains differ from real stains, we use computational metrics. Notably, these are *stain and tissue-specific* measures for evaluating our virtual stains. While one-size-fits-all image perceptual metrics like the Frechet Inception Distance (FID) exist [41], they are difficult to interpret. We advocate gaining more specific understanding of the ways in which Virtual Stains fail by applying domain-specific biomedical knowledge to the evaluation process. We have computational metrics of two forms: First, a domain-specific performance-validated DNN. We consider this to be an excellent standard for evaluation of generated images. The DNN produces Gleason grades, which is the clinically relevant output of the prostate H&E stain. In addition, while DNNs are notorious for out-of-set generalization issues [42], no such issues are encountered in the comparison of virtual versus real stains, despite the DNN not being trained on any virtual slides. This suggests that virtual stains are not out-of-distribution, while simultaneously demonstrating the fidelity of clinically-relevant information. The degree of agreement between virtual and real for this DNN exceed common kappa values of agreement between human pathologists substantially. For PIN-4 IHC, no such model for a clinically-relevant standard exists. Instead, we implement classical image segmentation techniques. We compare virtual versus real images to a control of serial tissue microarray sections, and find that for all domain-specific metrics, Virtual Stains are closer to their real counterparts than serial sections are to one another. Given that serial sections are functionally equivalent for the purpose of diagnostic use, we consider serial-section control experiments to be an excellent standard of quantitative analysis that we hope will be adopted more in future digital pathology validations.

In summary, we demonstrate both the feasibility of AF-based virtual stains of H&E and PIN-4 IHC for Prostate Cancer, as well as extensive qualitative and quantitative evaluation to exemplify a standard of rigor and clinical utility for digital pathology and virtual-staining technology research.

## Data Availability

The data for the patient samples cannot be shared under the restrictions placed by the institutional review board. The data storage and retrieval infrastructure was built and managed by Google LLC.

## Authors’ Contributions

P.F.W., A.H., D.F.S, L.R., J.R., P.C. contributed to study conception and design. P.F.W. led histopathology and image data generation, and pathologist annotation data workflows. C.S., M.G. led the development and calibration of the multispectral microscope and autofluorescence data collection. C.M., Y.W., J.P., K.N., P-H.C.C., T.J., E.W. wrote code for the data infrastructure, network architecture and training and testing pipelines of virtual stainer and scoring models. C.M., Y.W., J.P., T.J., and E.W. performed computational analysis of models. P.F.W. advised and reviewed model iterations and analyses.

## Data Availability

Data from patient samples cannot be shared under the restrictions placed by Verily’s contractual agreements with the tissue source.

## Code Availability

The pix2pix architecture is open sourced at and is available with tutorials at https://www.tensorflow.org/tutorials/generative/pix2pix. Releasing a trained binary or working code of our internal tooling, infrastructure and hardware is not feasible. However, we have described the neural network infrastructure in sufficient detail in Section : Materials and Methods to allow independent replication. All the algorithmic components of our work are all built on open source repositories: Python 3.6 packages Numpy, Scipy, OpenCV, Pandas, Seaborn and Matlplotlib were used for feature extraction, preprocessing, training and evaluation, statistical analysis and plotting. Tensorflow 2.0 with Keras was used to build, train and test the neural network models.

## Acknowledgements

The authors thank Leica Biosystems and Verily for supporting this collaborative work. We thank members of the translational pathology team for tissue processing, quality checking, staining, and imaging: Robert Findlater, Vanessa Velez, Julia Sigman, Hardik Patel, Tzu-Chien Wang. We thank members of program management and operations teams: Susan Kram, Nina Lottsfeldt, Janelle Chang-Clark, Fraser Tan, Robert Nagel, Allen Chai, and Craig Mermel. We thank members of Verily LIMS and lab engineering teams. We thank Dr. Trissia Brown and Dr. Isabelle Flament-Auvigne for giving us feedback on the quality of our virtual images. We thank Fabien Beckers for program leadership, Dr. Sudha Rao for pathology expertise, and Melissa Miao for partnership development.

## Ethics and Funding

Verily Life Sciences, LLC reports 3 pending patents on Virtual Staining and 1 on an AI-assisted Prostate Cancer Process.

P.F.W., C.M., Y.W., J.P., C.S., M.G., A.H., K.N., P-H.C.C., T.J., E.W., D.F.S, P.C. are current or former employees of Alphabet Inc. (Verily and Google) and performed work for this study during their tenure at Alphabet. They also report equity ownership in Alphabet at the time of work. L.R., J.R. are current or former employees of Danaher Corporation (Leica Biosystems) and performed work for this study during their tenure at Danaher.

