## Supplementary Materials for "Clinical-Grade Validation of an Autofluorescence Virtual Staining System with Human Experts and a Deep Learning System for Prostate Cancer"

Additional features of the dataset are provided. In Supplementary Table 1, we can see the number of slides sections and cases split into TRAIN, EVAL, and TEST for the purpose of machine learning and validation. In Supplementary Figure 1, we can see the distribution of different diagnostic categories among these splits, and verify that classes are relatively balanced between them. Splits were performed on the basis of patients, as we wish to train the model to generalize between patients, not just within patients.

| Split | Train | Eval | Test | Total |
|---|---|---|---|---|
| H&E Stained Sections | 188 | 188 | 423 | 799 |
| PIN-4 Stained Sections | 188 | 188 | 423 | 799 |
| Cases | 187 | 188 | 421 | 796 |

Supplementary Table 1. Number of unique slides and patients in the TRAIN/EVAL/TEST splits used to train and validate the virtual stainer models.

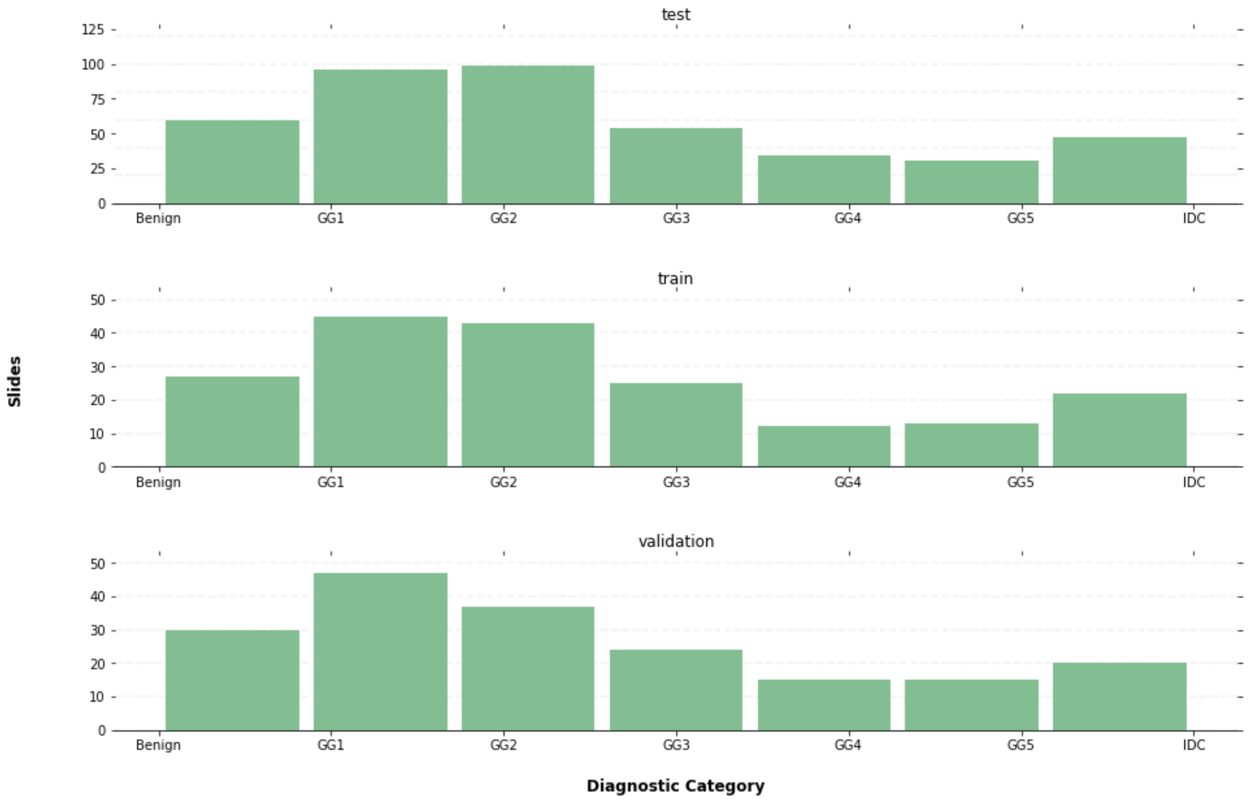

Supplementary Figure 1: Distributions of ground-truth diagnostic categories between each of the data splits for all sections. We can see that the randomly-chosen splits are relatively balanced with respect to diagnosis. This is a virtue of having a relatively large dataset.

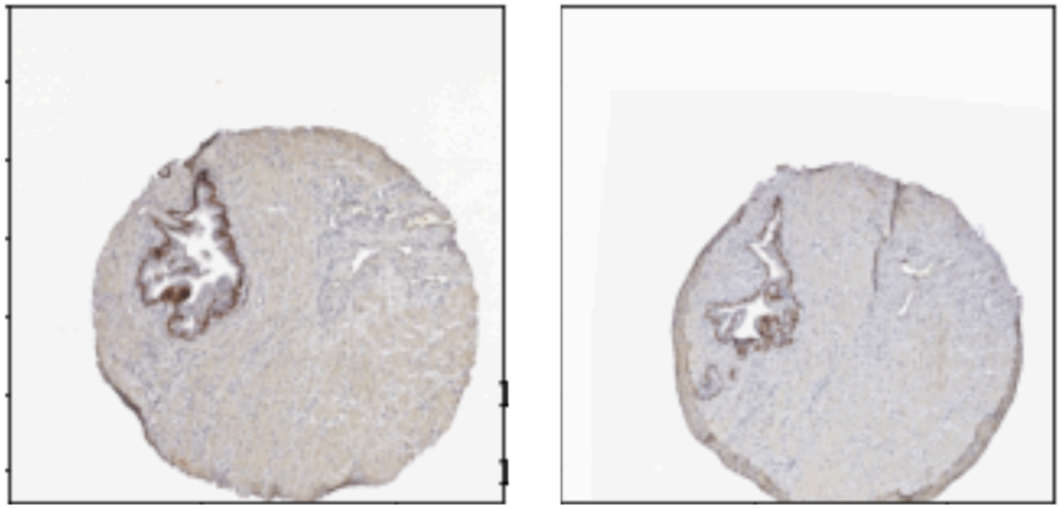

Supplementary Figure 2: An example of two serial PIN-4 Prostate TMA sections. We can see that the two sections are quite similar. Given their diagnostic equivalence, we use serial TMA sections like these as a control comparison for our computational image metrics.

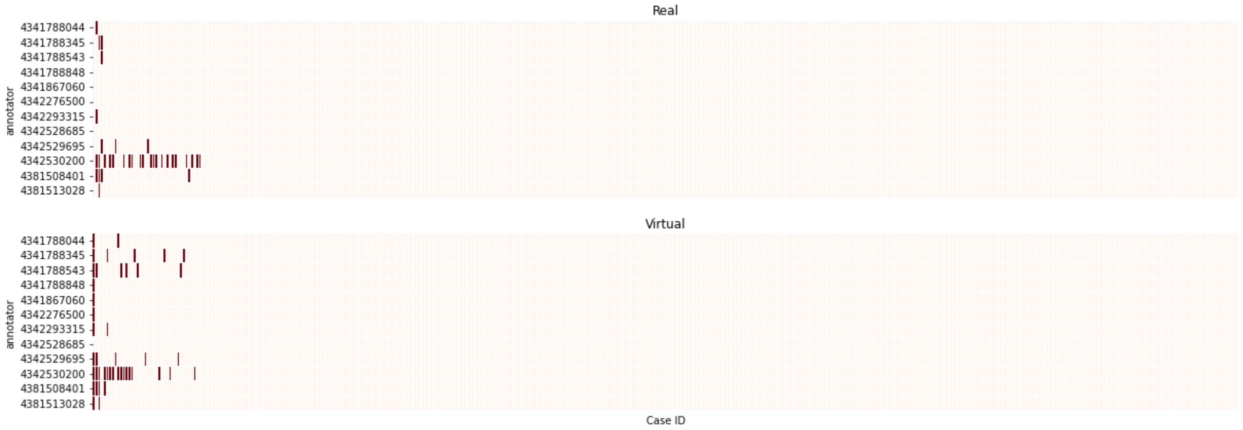


Supplementary Figure 3: Image Quality Review for H&E Images. Human Pathologists rated whether or not an image met their standard for quality to be able to give a confident diagnosis, for both real and virtual slides. No single slide was declared unusable by every pathologist. A total of 34 quality flags on real slides and 45 quality flags on virtual slides were made. (out of about 9500 reviews per set)

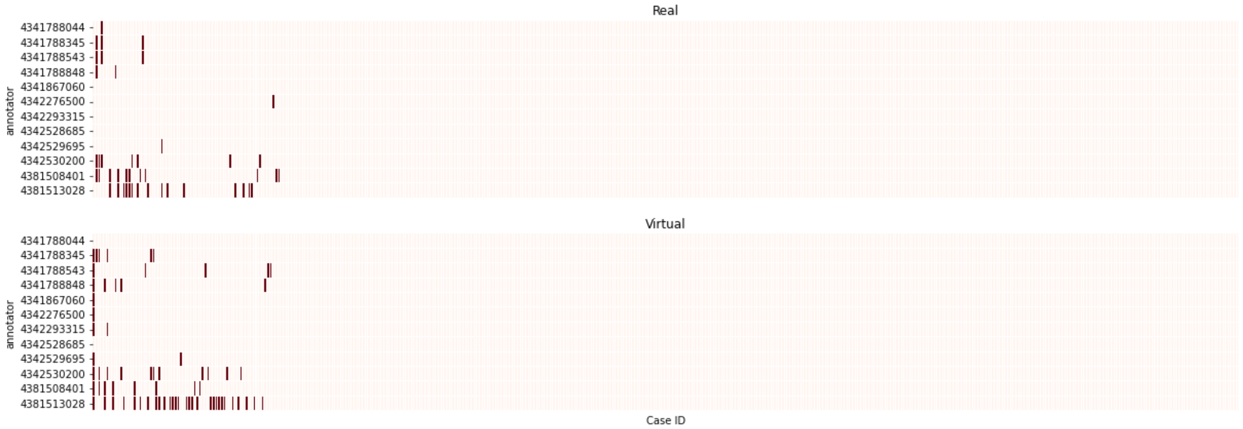


Supplementary Figure 4: Image Quality Review for PIN-4 Images. Human Pathologists rated whether or not an image met their standard for quality to be able to give a confident diagnosis, for both real and virtual PIN-4 slides. No single slide was declared unusable by every pathologist. A total of 44 quality flags on real slides and 70 quality flags on virtual slides were made. (out of about 9500 reviews per set)

### Supplementary Methods

#### Computational Metrics

Here we describe precisely the algorithmic metrics we use to compare real and virtual images.

##### Image Color Deconvolution

In order to extract meaningful biological information about the PIN-4 stains in an automated way, we use a color deconvolution technique, to separate the PIN-4 images into channels representing hematoxylin, AMACR, and Basal Stain components. Color deconvolution can be phrased as a non-negative matrix factorization problem: Consider an (raveled) image matrix $I$ of shape $(N, 3)$ representing $N$ pixels and 3 color channels. First, we transform this matrix into a set of values representing *absorbance* rather than *reflected light.* We do this because the amount of protein present, and therefore dye, should correspond to the absorbance of the material, not the intensity of reflected light collected by the imaging system.[37] The Beer-Lambert Law [38] gives us the relationship between these two:

$$I = I_0 e^{-A} \Rightarrow$$

$$A = -ln\left(\frac{I}{I_0}\right) \Rightarrow$$

$$A \propto -ln\left(\frac{I}{I_0}\right)$$

Given this absorbance matrix, we assert that the absorbance is the result of the concentration of relevant biomarker, multiplied by the color corresponding to that dye: $A = MC$, where $M$ is our deconvolved image, and $C$ is the “color absorbancy matrix”, representing the values of each color. This can be taken from the literature, or sampled from any PIN-4 image. At this point, we can use the Alternating-Least-Squares method (ALS) [39] to factorize *A,* with our sampled value for *C* taken as an initialization value.

#### PIN-4 Computational Image Metrics

Given deconvolved components representing nuclear (hematoxylin), AMACR (Fast Red), and basal cell markers (DAB), we can construct several metrics:

**AMACR Jaccard Distance:** On AMACR-positive patches, the signal occupies a relatively large fraction of tissue area. We should expect, between virtual stains vs. ground truth, or between serial TMAs, that the areas covered will be fairly similar. Given that we care more about the areas indicated by the stain than we care about the exact saturation of the AMACR, it is appropriate to classify image areas as “AMACR+” or negative, and compare them with Jaccard Distance. To do this, first we binarize the AMACR component of our deconvolved matrix *M* above with a simple Otsu Threshold. [40] Then, we take our two patches (whether corresponding Virtual/Real PIN-4 sections, or serial TMAs), and look at the Jaccard Distance, a value in between 0 and 1 corresponding to the amount of overlap between the two AMACR segmentation maps:

$$J_d(X_1, X_2) = \frac{X_1 \cap X_2}{X_1 \cup X_2}$$

**Nuclear and Basal Cell ROC Metrics:** For the small segments representing nuclear and basal cell signals, we use a similar approach to the AMACR analysis above. We use Otsu Thresholding to segment Basal Cells and Nuclear cells. The only difference is we add some classical image post-processing techniques to assert priors given that we know the approximate size of nuclei and Basal Cells. Given segments representing each, we can treat virtual staining like a signal detection problem. For each nuclei/basal cell in the ground truth, does it overlap with a cell in the predicted virtual image? If so, this is a true positive. If not, it is a false negative. Given these, as well as false positives and true negatives, we can count cells to get highly human-comprehensible precision and recall metrics.
